# Alpha-Band EEG Dynamics During Naturalistic Storytelling Interaction in Older Adult–Caregiver Dyads

**DOI:** 10.64898/2026.07.09.26357345

**Authors:** Supalak Khemthong, Winai Chatthong

**Affiliations:** Occupational Therapy Division, Faculty of Physical Therapy, Mahidol University, Phuttamonthon 4 Road, Salaya, Nakhon Pathom 73170, Thailand

**Keywords:** aging, alpha power, EEG, inter-brain similarity, naturalistic interaction, storytelling

## Abstract

Naturalistic social interaction provides an important context for examining how cognitive engagement is reflected in brain activity, yet electroencephalographic evidence from older adult–caregiver interaction remains limited. This study examined alpha-band EEG dynamics during museum-based storytelling interaction in older adult–caregiver dyads. Thirty-two dyads, comprising 32 older adults and 32 caregivers, completed cognitive and psychological screening and underwent EEG recording during eyes-closed resting, eyes-open resting, storytelling, and listening conditions. Relative alpha power was analyzed using a predefined 10-electrode sensor-level set covering frontal, frontotemporal, temporal, central, and parietal midline regions. Task-related alpha modulation was examined relative to eyes-open resting. Associations between cognitive performance and Cz alpha power were tested using MoCA scores, and dyad-level alpha-band inter-brain similarity was examined using spatial alpha-power patterns with within-site shuffled-dyad surrogate comparisons.

Alpha power was higher during eyes-closed resting and lower during storytelling and listening relative to eyes-open resting, indicating robust task-related modulation of alpha activity during naturalistic narrative interaction. Associations between MoCA scores and Cz alpha power were weak, condition-specific, and did not survive false discovery rate correction. During storytelling, dyad-level alpha-band inter-brain similarity was modestly higher than within-site shuffled-dyad estimates, but this effect did not remain significant after correction across conditions.

These findings suggest that alpha-band EEG activity is sensitive to naturalistic storytelling and listening in older adult–caregiver dyads. However, cognitive associations and dyad-level inter-brain similarity were modest and should be interpreted cautiously. The study demonstrates the feasibility of applying EEG to real-world social-cognitive interaction while highlighting the need for larger samples, behavioral coding, and time-resolved dyadic EEG methods to clarify mechanisms of interpersonal neural coordination.

## 1. Introduction

Naturalistic social interaction provides an important context for studying the relationship between cognitive engagement and physiological brain activity. Everyday communication requires attention, memory retrieval, semantic organization, affective expression, and partner-directed behavior. These processes are particularly relevant in later life, when cognitive functioning is shaped not only by individual neurocognitive status but also by social participation and meaningful interpersonal engagement.

Population aging has increased interest in identifying neurocognitive processes that support cognitive functioning in older adults. Mild cognitive impairment is commonly conceptualized as an intermediate state between typical cognitive aging and dementia, characterized by measurable cognitive difficulties while functional independence is largely preserved. Although clinical screening tools are useful for identifying cognitive vulnerability, they do not directly characterize the neural dynamics that support cognitive processing during everyday social interaction.

Lifestyle and social-contextual factors are increasingly recognized as relevant to cognitive aging. Epidemiological and prevention-oriented literature suggests that socially integrated and cognitively stimulating activities may be associated with lower dementia risk or slower cognitive decline (Fratiglioni et al., 2004; Livingston et al., 2020). However, the mechanisms through which socially meaningful activities engage attention, memory, and interpersonal coordination remain incompletely understood. This gap is important because many real-world cognitive activities, such as storytelling, require the integration of autobiographical memory retrieval, semantic organization, emotional expression, and partner-directed communication.

Electroencephalography provides a non-invasive method for examining neural oscillatory activity associated with cognitive processing. Cognitive impairment and neurodegenerative conditions have frequently been associated with altered spectral power, including increased slow-wave activity and reduced alpha rhythms, patterns sometimes described in prior literature as neural slowing (Healey & Kahana, 2020; Rondina et al., 2016, 2019). In addition to resting-state activity, oscillatory mechanisms are involved in attention, memory encoding, and task-related information processing. Alpha-band oscillations have been linked to attentional control and access to stored information (Klimesch, 2012), while event-related alpha/beta desynchronization has been interpreted as reflecting task-related cortical engagement (Pfurtscheller & Lopes da Silva, 1999). In aging research, altered alpha/beta desynchronization has also been associated with item–context binding difficulties in older adults (Karlsson & Sander, 2022), and hippocampal theta-gamma activity has been implicated in episodic memory formation (Griffiths et al., 2021). Age-related changes in alpha-beta desynchronization have also been associated with item-context binding difficulties in older adults (Karlsson & Sander, 2022). These findings suggest that alpha-band activity may provide a useful sensor-level index for examining attention- and cognition-related engagement during naturalistic narrative interaction.

Naturalistic social interaction adds another layer of complexity because cognitive processing unfolds jointly across interacting individuals. Hyperscanning and relational neuroscience frameworks emphasize that inter-brain findings should be interpreted in relation to behavior, social context, shared sensory input, task structure, and interpersonal factors (De Felice et al., 2025; Wolff & Dumas, 2026). Prior work shows that inter-brain coupling can occur during social interaction and cooperative tasks (Dumas et al., 2010; Kinreich et al., 2017), while naturalistic conversation studies suggest that synchrony may be associated with shared identity and social connectedness (Hinvest et al., 2025). Recent methodological work further indicates that dyadic social interaction may involve both temporal synchrony and spatial similarity across distributed brain networks (Li et al., 2026), and that network-based approaches may help clarify complex inter-brain dynamics beyond simple pairwise correlations (Kim et al., 2025). At the same time, spectral similarity between interacting individuals should not automatically be interpreted as socially meaningful neural synchrony, because similarity may also arise from common sensory input, shared task structure, turn-taking, or environmental context.

Despite these advances, naturalistic EEG studies of older adult–caregiver interaction remain limited. Museum-based storytelling provides a useful ecological paradigm because it combines visual prompts, autobiographical recall, attentional engagement, semantic organization, and interpersonal communication. Unlike laboratory tasks with constrained stimuli and responses, museum-based storytelling captures cognitive processing under more natural conditions while retaining sufficient procedural structure for EEG measurement. This setting also allows investigation of how older adults and caregivers engage with shared cultural and autobiographical material in a real-world social context.

The present study examined alpha-band EEG dynamics during museum-based storytelling interaction in older adult–caregiver dyads. Alpha-band activity was specified as the primary frequency band of interest, and relative alpha power was analyzed using a predefined 10-electrode sensor-level set to characterize task-related alpha modulation and dyad-level spatial similarity. Cz alpha power was retained as a focused central region of interest for cognition-related analyses because central alpha activity is theoretically relevant to attentional engagement and sensorimotor integration during storytelling and listening.

The study combined hypothesis-guided and exploratory aims. First, we examined whether alpha power differed across resting, storytelling, and listening conditions, using eyes-open resting as the within-participant baseline. Second, we examined whether Cz alpha power was associated with cognitive performance, measured using MoCA scores. Third, because inter-brain similarity during naturalistic older adult–caregiver storytelling remains underexplored, dyad-level alpha-band inter-brain similarity was treated as exploratory. We tested whether observed dyad-level alpha spatial similarity exceeded surrogate dyad estimates generated by shuffled pairings within the same museum site. This approach provided a cautious test of whether true dyads showed greater alpha-band similarity than would be expected from participants exposed to similar museum and task contexts but not interacting as a dyad.

## 2. Methods

### 2.1 Participants and recruitment

Thirty-two older adult–caregiver dyads, comprising 64 participants, were recruited from community visitors participating in museum-based storytelling programs in Thailand. Sixteen dyads were recruited from Museum Siam and 16 dyads were recruited from the National Science Museum. Older adult participants were aged 60 years or older and attended the activity with a caregiver, family member, or volunteer companion. Caregivers were aged 25–59 years. Recruitment was conducted through community outreach and museum education activities.

All participants provided written informed consent before participation. Participants were informed about the voluntary nature of the study, confidentiality of their data, and compensation for participation. The study received ethical approval from the XXX University Central Institutional Review Board.

### 2.2 Inclusion and exclusion criteria

Older adults were eligible if they were aged 60 years or older, legally competent, able to communicate in XXX, and able to use a mobile phone to take photographs during the museum activity. They were screened using the XXX version of the Patient Health Questionnaire-9 and were included if their scores indicated no to mild depressive symptoms. Cognitive status was screened using the Thai version of the Montreal Cognitive Assessment, and participants scoring above 16 were eligible, indicating cognitive performance ranging from normal cognition to possible mild cognitive impairment.

Caregivers were eligible if they were aged 25–59 years, legally competent, able to communicate in XXX, able to use a mobile phone to take photographs, and accompanying the older adult as a relative, family member, paid caregiver, or volunteer companion.

Participants were excluded if they had neurological disorders associated with severe cognitive impairment, including dementia, stroke, Parkinson’s disease, or delirium; moderate-to-severe psychiatric illness; active hallucinations; current medication or treatment known to substantially influence EEG recordings; or abnormal involuntary movements that could interfere with EEG acquisition.

### 2.3 Study setting and storytelling task

Data collection was conducted in two museum environments designed to support culturally meaningful storytelling and intergenerational engagement. The storytelling task used a photo-elicitation approach. Before EEG recording, each dyad visited six exhibition rooms in the museum for approximately 60 minutes. During the visit, older adults and caregivers were invited to take photographs of museum objects, scenes, or exhibition spaces that they found personally meaningful or emotionally salient. Participants could take photographs independently or take photographs for one another.

After the museum visit, each participant selected one photograph that was most personally meaningful. This selected photograph was used as the prompt for the storytelling task. Participants were asked to prepare a brief narrative guided by three questions: (1) What is shown in the photograph? (2) Why was this photograph meaningful or impressive? and (3) How does this image relate to health, well-being, or happiness in later life?

The storytelling task followed a structured but naturalistic protocol. During the EEG-recorded interaction, the older adult first narrated the selected photograph while the caregiver listened. The caregiver was allowed to respond naturally, ask clarifying questions, or share related reflections without leading the narrative. The caregiver then became the storyteller, and the older adult became the listener. This role-switching structure allowed both members of the dyad to participate in storytelling and listening conditions.

The protocol was designed to balance ecological validity with reproducibility. Narrative content was not identical across dyads because the study aimed to capture naturalistic storytelling; however, the task structure, prompts, EEG timing, and recording procedures were standardized across sites.

### 2.4 Experimental procedure

The experimental procedure consisted of six sequential phases: participant enrolment, museum visit, photograph selection, EEG recording, post-recording care, and data analysis.

**FIGURE 1.**
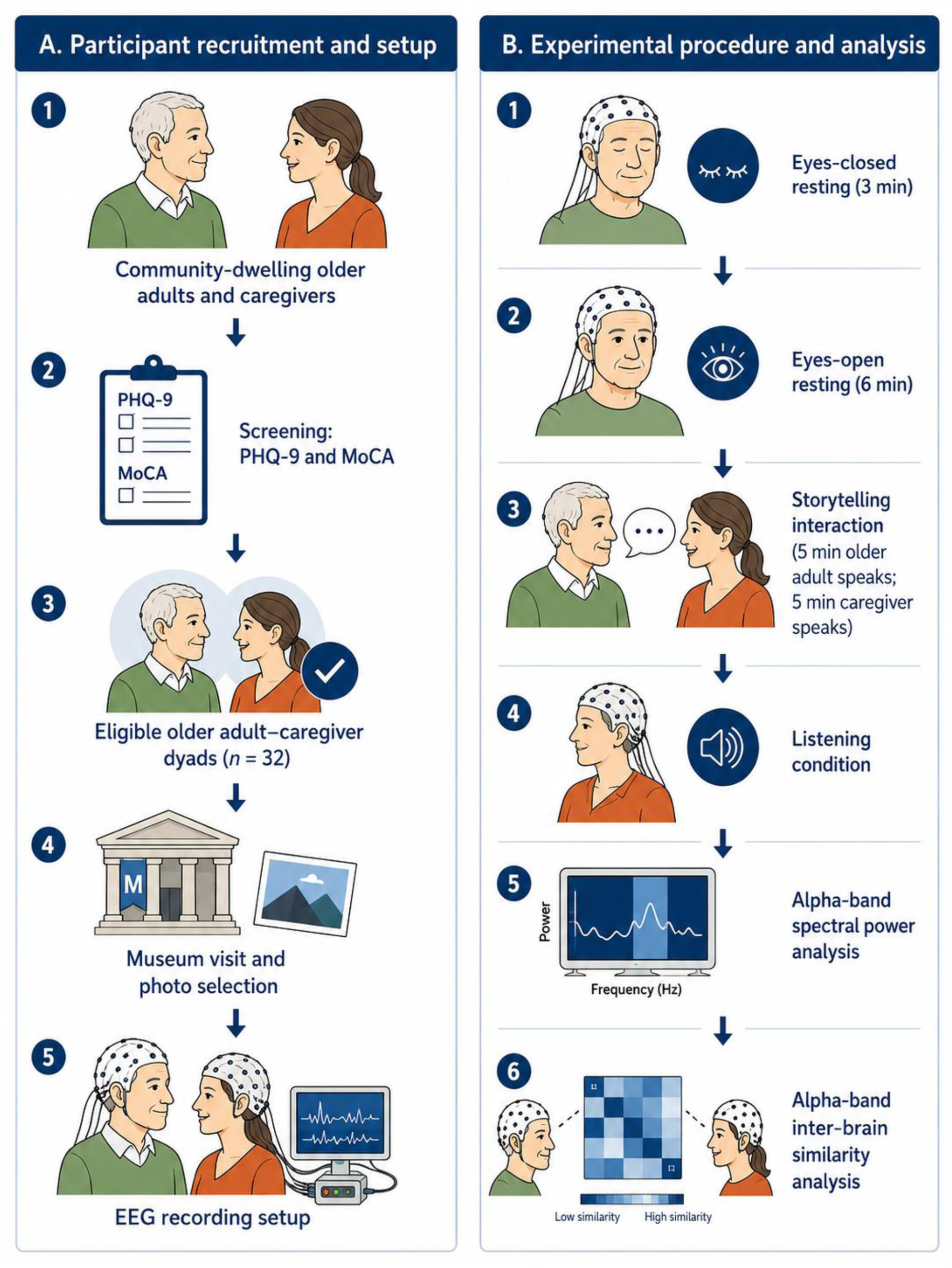
Experimental procedure and EEG analysis overview.

First, participants completed the enrolment process. Demographic and background information was collected, including age, sex, education, socioeconomic background, health history, caregiver relationship, and museum visit history. Cognitive and psychological screening was then conducted using the MoCA and PHQ-9. Written informed consent was obtained from all participants.

Second, each dyad visited six exhibition rooms in the museum for approximately 60 minutes and took photographs of personally meaningful museum objects, scenes, or exhibition spaces.

Third, after the museum visit, each participant selected one photograph to serve as the prompt for the storytelling interaction.

Fourth, EEG recording was conducted while participants wore the EEG cap. EEG setup and signal preparation lasted approximately 11 minutes. EEG data were then recorded under resting and interaction conditions. Eyes-closed resting EEG was recorded for 3 minutes, followed by eyes-open resting EEG for 6 minutes while dyad members sat facing one another without storytelling. The storytelling interaction was then recorded for 10 minutes, consisting of 5 minutes in which the older adult narrated the selected photograph while the caregiver listened, followed by 5 minutes in which the caregiver narrated while the older adult listened.

Fifth, after EEG recording, participants received post-recording care. Conductive gel was removed from the hair and scalp using dry and wet tissues to ensure participant comfort.

Finally, EEG data were processed for spectral analysis and topographical mapping. Photo and narrative materials were used to contextualize the storytelling interaction but were not treated as fixed experimental stimuli.

2.5 Cognitive and psychological measures

Cognitive performance was assessed using the XXX version of the Montreal Cognitive Assessment. The MoCA evaluates visuospatial and executive function, attention, memory, language, abstraction, and orientation. Depressive symptoms were assessed using the XXX version of the Patient Health Questionnaire-9, a nine-item self-report measure scored from 0 to 27. These measures were used for screening and participant characterization rather than for clinical diagnosis. MoCA scores were also used as a continuous index of cognitive performance in correlation analyses.

### 2.6 EEG recording and preprocessing

EEG signals were recorded using a BrainMaster Discovery 24E QEEG system (K150498; BrainMaster Technologies, Inc.). The system included a 19-position electrode cap arranged according to the international 10–20 system and was connected to a 12-channel signal acquisition unit. Recordings were obtained during eyes-closed resting, eyes-open resting, storytelling, and listening conditions using the same hardware and montage settings.

Before recording, electrode impedance was checked and maintained below 5 kΩ for all channels, with inter-electrode impedance differences kept within 2 kΩ. EEG data were sampled at 250 Hz and referenced to linked ears. Continuous EEG signals were notch filtered at 50 Hz to reduce line-noise contamination and bandpass filtered from 1 to 30 Hz before spectral analysis.

EEG preprocessing and spectral analysis were conducted offline using NeuroGuide Deluxe QEEG software, version 3.0.7.3. Automated artifact detection was applied using an amplitude threshold of ±100 µV to exclude segments containing gross artifacts, including eye blinks, swallowing, chewing, head movement, and body movement. Artifact sensitivity was set to high to reduce contamination from drowsiness-related activity and eye movements. Amplitude selection was adjusted using the multiplier method, with values set between 0.9 and 1.0.

Independent component analysis was applied to identify and remove components associated with ocular and muscle artifacts. Automated preprocessing was followed by manual visual inspection to ensure that retained segments were suitable for spectral analysis and topographical mapping. A minimum of 60 seconds of artifact-free EEG data was retained for each analyzed condition.

Power spectral density was estimated using Welch’s method with 2-second epochs and 50% overlap. NeuroGuide generated standard spectral estimates across delta, theta, alpha, and beta frequency bands. In the inferential analysis, alpha-band activity was specified as the primary frequency band of interest to reduce the number of statistical comparisons and to focus on cognition-related oscillatory activity. Relative alpha power was calculated as alpha power divided by total power across the 1–30 Hz range.

Topographical maps were generated as descriptive visualizations of sensor-level alpha-power patterns across resting, storytelling, and listening conditions. Z-score brain maps derived from the NeuroGuide normative database were used descriptively and were not interpreted as clinical diagnostic maps or evidence of localized neural generators. Individual alpha peak frequency was not controlled for in the present analysis.

### 2.7 Alpha-band inter-brain similarity analysis

Alpha-band inter-brain similarity was defined as dyad-level similarity in sensor-level alpha-power patterns between an older adult and their caregiver during the same experimental condition. This term was used instead of stronger claims about neural synchrony because the present analysis was based on spatial similarity in spectral power rather than continuous time-resolved hyperscanning connectivity.

For each dyad and condition, relative alpha power values were extracted from a predefined 10-electrode sensor-level set: Fp1, Fp2, F3, F4, F8, T3, T4, Fz, Cz, and Pz. These values were arranged as spatial alpha-power patterns separately for the older adult and caregiver. Inter-brain similarity was calculated using Pearson’s spatial correlation between the older adult’s and caregiver’s 10-electrode alpha-power patterns within each dyad. Correlation coefficients were transformed using Fisher’s z for statistical reporting. Higher Fisher’s z values indicated greater similarity in alpha-power distribution between dyad members.

To address the possibility that similarity was driven by shared task structure, common sensory input, museum environment, or procedural artifacts, a surrogate-dyad robustness check was conducted. Older adults and caregivers were randomly re-paired within the same museum site, thereby preserving site-specific museum context while disrupting the original dyadic pairing. Alpha-band inter-brain similarity was recalculated for each shuffled pairing across 10,000 permutations to generate a surrogate distribution of chance-level dyadic similarity. Observed dyad-level similarity was then compared with the surrogate distribution to determine whether true dyads showed greater alpha-band spatial similarity than would be expected from participants exposed to broadly similar museum and task contexts but not interacting as a dyad. The resulting permutation p value was calculated as the proportion of surrogate similarity values equal to or greater than the observed similarity value.

### 2.8 Statistical analysis

Statistical analyses were conducted using IBM SPSS Statistics. Participant characteristics were summarized using means and standard deviations for continuous variables and counts and percentages for categorical variables.

Alpha-band activity was specified as the primary frequency band of interest. To reduce the number of statistical comparisons and avoid post hoc interpretation of individual electrode findings, inferential EEG analyses were restricted to the predefined 10-electrode sensor-level alpha set. This set provided coverage of frontal, frontotemporal, temporal, central, and parietal midline regions relevant to attentional engagement, narrative processing, and social-cognitive task participation.

To evaluate task-related alpha modulation, alpha power was analyzed across four conditions: eyes-closed resting, eyes-open resting, storytelling, and listening. The eyes-open resting condition was used as the within-participant baseline to reduce inter-individual variability and to control for visual activation effects. For each participant, task-related difference scores were calculated as:

Δ Alpha Power = Relative Alpha Power Condition − Relative Alpha Power Eyes Open

Difference scores were summarized as mean differences, standard deviations, and 95% confidence intervals by participant role and condition.

Before conducting inferential analyses, normality of Δ Alpha Power values was assessed using the Shapiro–Wilk test. When Δ Alpha Power values were normally distributed, paired-samples t-tests were used to examine within-participant differences between eyes-open resting and eyes-closed resting, storytelling, or listening conditions. When normality assumptions were not met, Wilcoxon signed-rank tests were used. Effect sizes were reported using Cohen’s dz for paired-samples t-tests and rank-based effect sizes for nonparametric comparisons where appropriate.

To address multiple comparisons, p values from planned condition-based comparisons were adjusted using the Benjamini–Hochberg false discovery rate procedure. Both uncorrected p values and FDR-adjusted q values were reported. Findings that remained significant after FDR correction were interpreted as statistically robust within the exploratory framework, whereas findings that did not survive correction were interpreted cautiously as hypothesis-generating.

Associations between alpha power and cognitive or psychological measures were examined using correlation analyses. Cz alpha power was retained as a focused central region-of-interest analysis for cognition-related associations because central alpha activity was considered theoretically relevant to attentional engagement and sensorimotor integration during storytelling and listening. Normality of the relevant EEG and behavioral variables was assessed using the Shapiro–Wilk test before correlation analysis. Pearson’s correlation coefficients were used when both variables were normally distributed, whereas Spearman’s rank correlation coefficients were used when at least one variable was not normally distributed. Correlations were examined between relative alpha power and MoCA scores. Correlation coefficients, p values, FDR-adjusted q values, and 95% confidence intervals were reported.

Alpha-band inter-brain similarity analyses were conducted using Fisher’s z-transformed Pearson spatial correlations derived from the 10-electrode alpha-power patterns. Within-site surrogate-dyad analyses were used to evaluate whether observed dyad-level similarity exceeded chance pairing while preserving museum-site context. Permutation p values were reported, and FDR correction was applied across condition-specific surrogate comparisons.

A post hoc sensitivity analysis was conducted using G*Power 3.1 to estimate the minimum detectable effect size for participant-level and dyad-level analyses. Given the modest sample size and exploratory dyadic design, small effects were interpreted cautiously.

## 3. Results

### 3.1 Participant characteristics

Participant characteristics are summarized in Table 1. The final sample included 32 older adult–caregiver dyads, comprising 64 participants. Sixteen dyads were recruited from XXX and 16 dyads were recruited from the XXX Museum. The sample included 32 older adults and 32 caregivers, with equal representation across the two museum sites.

Older adults were older than caregivers, as expected from the dyadic design. PHQ-9 scores were low across both groups, indicating minimal depressive symptoms overall. MoCA scores suggested that cognitive performance among older adults ranged from normal cognition to possible mild cognitive impairment, whereas caregivers generally showed higher cognitive screening scores. Additional demographic characteristics, including sex, education, socioeconomic background, pre-existing conditions, museum visit history, and caregiver relationship, were summarized to clarify the sample and potential generalizability limits.

Across the total sample, the most common education levels were bachelor’s degree and postgraduate education. Most participants reported having sufficient income with savings or sufficient income for daily expenses. Pre-existing health conditions were reported by approximately half of the sample. Among caregivers, relationship roles included volunteer caregivers, adult children, paid caregivers, grandchildren, and other relatives.

Because participants were recruited through museum-based activities, the sample may represent older adults and caregivers who were sufficiently mobile, socially engaged, and willing to participate in cultural and narrative tasks. This recruitment context is considered in the Limitations section.

### 3.2 Task-related alpha-band EEG modulation

Topographical maps of relative alpha power across the predefined 10-electrode sensor-level set are presented in Figures 2. These maps were used descriptively to visualize spatial patterns of alpha-band activity across eyes-closed resting, eyes-open resting, storytelling, and listening conditions. Interpretation was restricted to sensor-level alpha-power patterns and was not extended to claims about specific cortical generators or clinical diagnostic maps.

**FIGURE 2.**
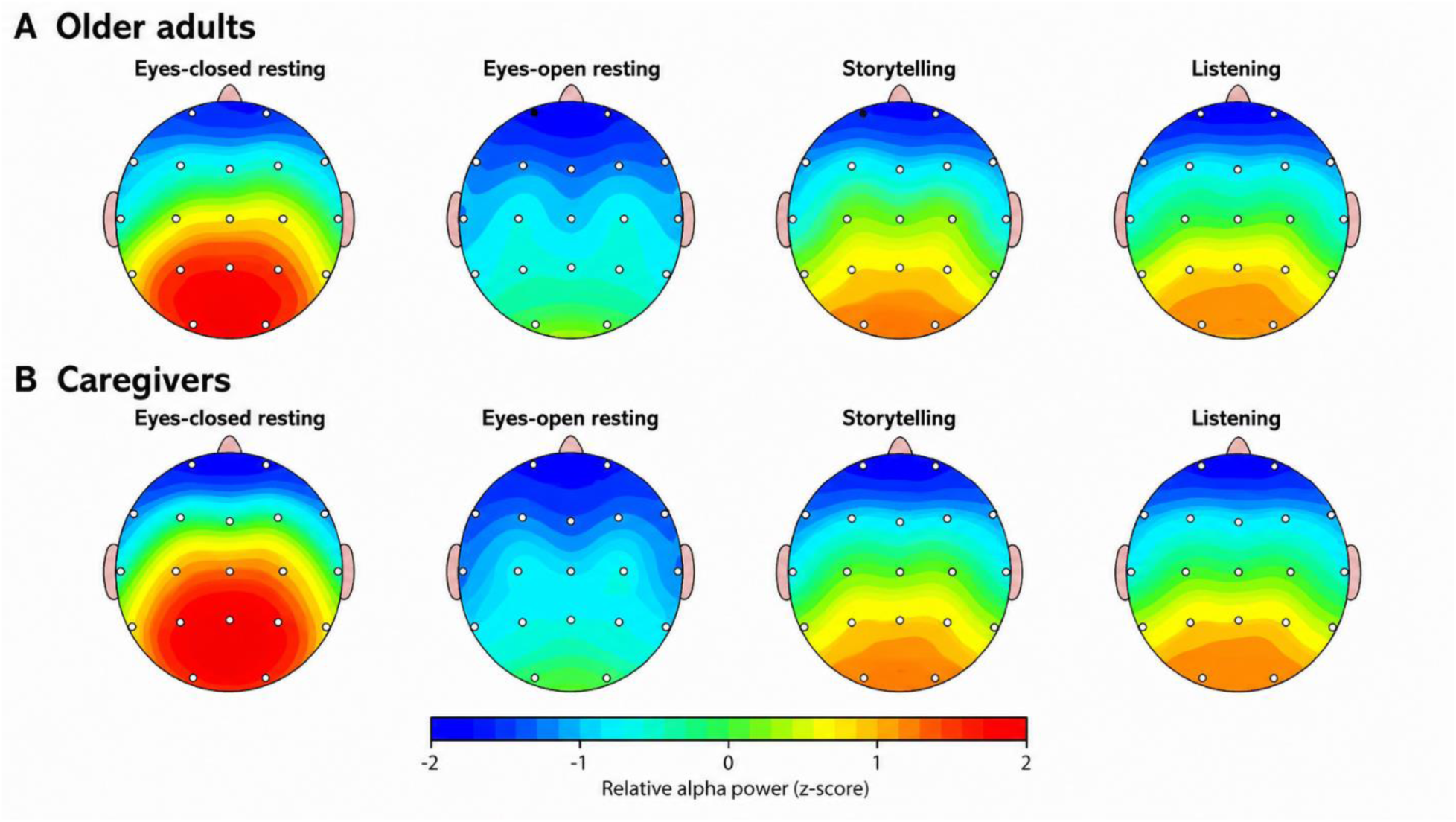
Alpha-band topographical maps across conditions and roles.

Formal condition-based comparisons are reported in Table 2. The eyes-open resting condition was used as the within-participant baseline. Across the 10-electrode alpha set, both older adults and caregivers showed higher alpha power during eyes-closed resting relative to eyes-open resting. In contrast, alpha power was lower during storytelling and listening relative to eyes-open resting. These effects remained significant after false discovery rate correction.

Among older adults, mean alpha power increased from 12.04 ± 4.65 during eyes-open resting to 23.05 ± 7.27 during eyes-closed resting, t(31) = 7.96, p < .001, q < .001, Cohen’s dz = 1.41. Alpha power decreased during storytelling, 7.77 ± 2.85, relative to eyes-open resting, t(31) = −6.41, p < .001, q < .001, Cohen’s dz = −1.13. A similar reduction was observed during listening, 7.80 ± 3.53, t(31) = −6.24, p < .001, q < .001, Cohen’s dz = −1.10.

Among caregivers, mean alpha power increased from 11.05 ± 4.87 during eyes-open resting to 30.16 ± 12.96 during eyes-closed resting, t(31) = 8.72, p < .001, q < .001, Cohen’s dz = 1.54. Alpha power decreased during storytelling, 7.29 ± 2.17, relative to eyes-open resting, Wilcoxon W = 41, p < .001, q < .001, rank-based effect size r = −0.81. Alpha power also decreased during listening, 7.59 ± 3.19, Wilcoxon W = 15, p < .001, q < .001, rank-based effect size r = −0.96.

Together, these findings indicate robust task-related modulation of alpha-band EEG activity. Eyes-closed resting was associated with increased alpha power, whereas storytelling and listening were associated with alpha suppression relative to eyes-open resting.

### 3.3 Associations between central alpha activity and cognitive performance

Cz alpha power was examined as a focused central region-of-interest analysis for cognition-related associations because central alpha activity was considered theoretically relevant to attentional engagement and sensorimotor integration during storytelling and listening. The relationship between cognitive performance and central alpha activity was examined by correlating MoCA scores with relative alpha power at Cz across experimental conditions. Full statistical results are reported in Table 3. Because MoCA scores were not normally distributed, Spearman’s rank correlation coefficients were used for these analyses.

Across conditions, associations between MoCA scores and Cz alpha power were small and variable. No association remained statistically significant after false discovery rate correction across the four condition-specific Cz alpha–MoCA correlations. During eyes-open resting, MoCA scores showed a weak negative association with Cz alpha power at the uncorrected level; however, this association did not survive FDR correction. Associations during eyes-closed resting, storytelling, and listening were weak and non-significant.

These findings indicate that the present data do not provide robust evidence for a consistent relationship between MoCA scores and Cz alpha power. Therefore, condition-specific alpha–MoCA findings were interpreted cautiously as exploratory cognition-related associations and should not be considered evidence of a diagnostic biomarker.

### 3.4 Alpha-band inter-brain similarity

Dyad-level alpha-band inter-brain similarity was examined using relative alpha power from the predefined 10-electrode sensor-level set. For each dyad and condition, alpha power values from Fp1, Fp2, F3, F4, F8, T3, T4, Fz, Cz, and Pz were arranged as spatial alpha-power patterns separately for the older adult and caregiver. Pearson spatial correlations were calculated between dyad members and transformed using Fisher’s z. Higher Fisher’s z values indicated greater similarity in alpha-power distribution between dyad members.

Across all dyads (Table 4), alpha-band inter-brain similarity during storytelling showed a mean Fisher’s z of 1.00 ± 0.48. Relative to eyes-open resting, storytelling showed a small increase in inter-brain similarity, but this difference did not reach statistical significance after FDR correction, t(31) = 1.90, p = .067, q = .199. Listening also showed a small, non-significant increase relative to eyes-open resting, t(31) = 1.12, p = .272, q = .272. Eyes-closed resting showed lower inter-brain similarity than eyes-open resting, but this difference was also not significant, t(31) = −1.54, p = .133, q = .199.

To evaluate whether observed dyad-level similarity exceeded chance pairing while accounting for museum context, a within-site surrogate-dyad analysis was conducted. Older adults and caregivers were randomly re-paired within the same museum site across 10,000 permutations, thereby preserving site-specific museum context while disrupting the original dyadic pairing. During storytelling, observed dyad-level similarity was slightly higher than the surrogate distribution, observed mean Fisher’s z = 1.00, surrogate mean = 0.93, one-tailed permutation p = .041. However, this effect did not remain significant after FDR correction across conditions, q = .092. Similar within-site surrogate analyses for eyes-closed resting, eyes-open resting, and listening also did not survive FDR correction.

Site-stratified exploratory checks suggested variability across museum settings, but no site-specific surrogate comparison survived FDR correction. Therefore, the present findings are best interpreted as exploratory evidence of alpha-band inter-brain similarity rather than definitive evidence of socially specific neural synchrony. The results suggest that true dyads showed modestly greater storytelling-related alpha-pattern similarity than shuffled dyads, but this effect was not sufficiently robust after correction for multiple comparisons.

## 4. Discussion

The present study examined alpha-band EEG dynamics during naturalistic museum-based storytelling interaction in older adult–caregiver dyads. The study combined hypothesis-guided analyses of task-related alpha modulation and cognition-related central alpha activity with exploratory analyses of dyad-level alpha-band inter-brain similarity. Three main findings emerged. First, alpha power showed robust task-related modulation: alpha power increased during eyes-closed resting relative to eyes-open resting and decreased during storytelling and listening relative to eyes-open resting. Second, associations between MoCA scores and Cz alpha power were weak, condition-specific, and did not survive false discovery rate correction. Third, dyad-level alpha-band inter-brain similarity during storytelling showed a modest elevation relative to within-site shuffled dyads, but this effect did not remain significant after correction across conditions. Together, these findings suggest that alpha-band EEG activity is sensitive to naturalistic storytelling and listening, while cognitive associations and dyadic similarity effects should be interpreted cautiously.

The reduction in alpha power during storytelling and listening relative to eyes-open resting is consistent with the view that alpha-band activity is sensitive to attentional engagement and task-related information processing. Alpha suppression during active cognitive tasks has commonly been interpreted as reflecting increased cortical engagement, semantic processing, or task-related information processing relative to resting or less demanding conditions (Klimesch, 2012; Pfurtscheller & Lopes da Silva, 1999). Storytelling requires autobiographical memory retrieval, semantic organization, affective expression, and partner-directed communication, while listening requires sustained attention, monitoring of narrative content, and interpersonal responsiveness. Prior work on episodic and narrative memory has also implicated oscillatory mechanisms in memory encoding, retrieval, and semantic elaboration (Griffiths et al., 2021; Healey & Kahana, 2020). In this context, alpha suppression may reflect increased cognitive engagement during active narrative processing relative to passive eyes-open resting. However, because the present study used sensor-level EEG rather than source-localized EEG, these findings should not be interpreted as evidence of specific cortical generators.

The cognition-related findings were more limited. Cz alpha power was retained as a focused central region of interest because central alpha activity is theoretically relevant to attentional engagement and sensorimotor integration during storytelling and listening. Prior EEG studies of cognitive aging have reported associations between cognitive performance and altered spectral power, including reduced alpha activity and increased slow-wave activity in aging or cognitive impairment contexts (Chino et al., 2024; Healey & Kahana, 2020; Rondina et al., 2016, 2019). However, the present data did not provide robust evidence for a consistent association between MoCA scores and Cz alpha power. Although a weak uncorrected association was observed during eyes-open resting, no condition-specific alpha–MoCA association remained significant after FDR correction. These findings suggest that Cz alpha power should not be interpreted as a diagnostic marker of cognitive status in the present sample. Rather, the alpha–MoCA analysis should be viewed as exploratory evidence that central alpha activity may relate weakly and variably to cognitive performance under some conditions.

The inter-brain findings also require cautious interpretation. Dyadic neural similarity was examined using alpha-band spatial patterns from a predefined 10-electrode sensor-level set rather than relying on a single electrode. This approach provided broader spatial coverage and reduced the risk of interpreting an isolated electrode-level association as dyadic neural coupling. During storytelling, observed dyad-level alpha similarity was modestly higher than the within-site surrogate distribution, suggesting that true dyads may show slightly greater alpha-pattern similarity than randomly re-paired participants from the same museum context. However, this effect did not survive FDR correction across conditions. Therefore, the present findings are best described as exploratory alpha-band inter-brain similarity rather than definitive evidence of socially specific neural synchrony.

This cautious interpretation is important for psychophysiological research on naturalistic interaction. Hyperscanning and relational neuroscience studies show that inter-brain measures may be associated with shared attention, interpersonal coordination, social connectedness, and cooperative interaction (De Felice et al., 2025; Dumas et al., 2010; Hinvest et al., 2025; Kinreich et al., 2017; Wolff & Dumas, 2026). At the same time, inter-brain similarity can also be influenced by shared sensory input, environmental context, task timing, speech structure, and turn-taking. Recent methodological discussions emphasize that dyadic neural measures should be interpreted in relation to behavioral coordination, task structure, and appropriate control or surrogate comparisons (Kim et al., 2025; Li et al., 2026; Wolff & Dumas, 2026). In the present study, older adults and caregivers experienced the same museum environment and participated in a shared storytelling protocol. The within-site surrogate analysis helped control for museum context, but the study did not fully model speech timing, gaze, turn-taking dynamics, or moment-to-moment behavioral coordination. Thus, the observed alpha similarity may reflect a combination of dyad-specific engagement, shared task structure, and common environmental context.

The study contributes to psychophysiological research by demonstrating the feasibility of recording and analyzing EEG during a structured but naturalistic social-cognitive task in older adult–caregiver dyads. Many EEG studies of cognitive aging rely on resting-state recordings or highly controlled laboratory tasks, which may not fully capture how cognitive processing unfolds during everyday social interaction (Chino et al., 2024; Healey & Kahana, 2020). The present paradigm extends this work by examining alpha-band activity during real-world narrative interaction, where cognitive processing unfolds through memory retrieval, meaning-making, and interpersonal communication. Museum-based and other naturalistic contexts may be particularly useful for studying social engagement because they combine sensory stimulation, autobiographical reflection, and interpersonal exchange in ecologically meaningful settings (Dikker et al., 2021). The findings support the value of naturalistic paradigms for studying brain–behavior relationships while also highlighting the methodological challenges of interpreting sensor-level EEG and dyad-level similarity measures outside the laboratory.

### 4.1 Limitations

Several limitations should be considered. First, the sample size was modest, particularly for dyad-level analyses. The post hoc sensitivity analysis indicated that the study was better powered to detect moderate effects than small effects. Therefore, weak or null findings should not be interpreted as definitive evidence of absence. Second, participants were recruited from museum-based activities, which may introduce sampling bias. Older adults and caregivers who attend museums or community education activities may be more mobile, socially engaged, and comfortable with cultural participation than the broader older adult population. This limits the generalizability of the findings to less mobile, socially isolated, or more clinically impaired older adults.

Third, EEG provides useful temporal information but limited spatial resolution. Therefore, the present sensor-level alpha findings should not be interpreted as evidence of localized neural generators. Fourth, individual alpha peak frequency was not controlled for, and fixed alpha-band definitions may have introduced inter-individual variability in alpha estimates. Fifth, alpha-band inter-brain similarity was examined using spatial spectral similarity rather than continuous time-resolved hyperscanning connectivity. Although a within-site shuffled-dyad analysis was added, speech timing, gaze, turn-taking, and shared sensory input were not fully modeled. Sixth, the cross-sectional design does not allow conclusions about whether storytelling engagement changes cognitive performance over time.

Future studies should include larger samples, preregistered hypotheses, time-resolved dual-brain EEG or hyperscanning methods, behavioral coding of interaction quality, and longitudinal follow-up. Future work should also examine whether museum site, caregiver relationship, familiarity, conversational quality, and cognitive-status differences moderate alpha-band inter-brain similarity.

### 4.2 Conclusion

This study provides evidence that alpha-band EEG activity is modulated during naturalistic storytelling and listening in older adult–caregiver dyads. Alpha power decreased during storytelling and listening relative to eyes-open resting, suggesting increased task-related engagement. However, associations between Cz alpha power and MoCA scores were weak and did not survive correction for multiple comparisons. Dyad-level alpha-band inter-brain similarity during storytelling showed a modest trend relative to within-site shuffled dyads, but this effect was not robust after FDR correction. Overall, the findings support the feasibility of using EEG in ecologically valid social contexts while emphasizing the need for cautious interpretation, stronger dyadic controls, behavioral coding, and time-resolved methods in future psychophysiological research.

## Data Availability

All data produced in the present study are available upon reasonable request to the authors.

## Declarations

### Ethical approval

This experimental study involving human participants received ethical approval from the XXX University Central Institutional Review Board XXX, Certificate of Approval No. XXX.

