## Supplementary material for "Alpha-Band EEG Dynamics During Naturalistic Storytelling Interaction in Older Adult–Caregiver Dyads": Table 1-4

**TABLE 1**. Participant characteristics by role and museum site.

| Group | n | Age, years | PHQ-9 | MoCA | Female, n (%) | Male, n (%) |
|---|---|---|---|---|---|---|
| Total sample | 64 | 56.70 ± 14.92 | 1.69 ± 2.20 | 25.69 ± 3.33 | 49 (76.6%) | 15 (23.4%) |
| Caregivers, total | 32 | 44.03 ± 9.43 | 2.16 ± 2.76 | 26.84 ± 2.03 | 26 (81.2%) | 6 (18.8%) |
| Older adults, total | 32 | 69.38 ± 5.68 | 1.22 ± 1.31 | 24.53 ± 3.95 | 23 (71.9%) | 9 (28.1%) |
| Caregivers – XXX | 16 | 49.62 ± 5.16 | 1.50 ± 1.10 | 27.50 ± 1.71 | 15 (93.8%) | 1 (6.2%) |
| Older adults – XXX | 16 | 72.31 ± 6.24 | 1.38 ± 1.36 | 24.38 ± 3.96 | 12 (75.0%) | 4 (25.0%) |
| Caregivers – XXX | 16 | 38.44 ± 9.51 | 2.81 ± 3.69 | 26.19 ± 2.17 | 11 (68.8%) | 5 (31.2%) |
| Older adults – XXX | 16 | 66.44 ± 3.05 | 1.06 ± 1.29 | 24.69 ± 4.06 | 11 (68.8%) | 5 (31.2%) |

*Note:* Values are mean ± standard deviation unless otherwise stated. PHQ-9 = Patient Health Questionnaire-9; MoCA = Montreal Cognitive Assessment. Museum XXX and XXX Museum each contributed 16 dyads.

Additional demographic characteristics may be reported in the text or supplementary material: bachelor’s degree, 31/64 (48.4%); postgraduate education, 21/64 (32.8%); sufficient income with savings, 37/64 (57.8%); sufficient income for daily expenses, 21/64 (32.8%); pre-existing health conditions, 33/64 (51.6%). Caregiver relationship roles included volunteer caregivers, 13/32 (40.6%); adult children, 10/32 (31.2%); paid caregivers, 4/32 (12.5%); grandchildren, 3/32 (9.4%); and other relatives, 2/32 (6.2%).

**TABLE 2.** Task-related comparisons of mean alpha power across the predefined 10-electrode set.

| Role | Comparison | Baseline EO, M (SD) | Condition, M (SD) | Δ Mean [95% CI] | Test statistic | Effect size |
|---|---|---|---|---|---|---|
| Older adults | EC vs EO | 12.04 (4.65) | 23.05 (7.27) | 11.01 [8.19, 13.84] | t(31) = 7.96** | dz = 1.41 |
| Older adults | EX vs EO | 12.04 (4.65) | 7.77 (2.85) | −4.27 [−5.62, −2.91] | t(31) = −6.41** | dz = −1.13 |
| Older adults | RE vs EO | 12.04 (4.65) | 7.80 (3.53) | −4.24 [−5.62, −2.85] | t(31) = −6.24** | dz = −1.10 |
| Caregivers | EC vs EO | 11.05 (4.87) | 30.16 (12.96) | 19.11 [14.64, 23.58] | t(31) = 8.72** | dz = 1.54 |
| Caregivers | EX vs EO | 11.05 (4.87) | 7.29 (2.17) | −3.76 [−5.37, −2.15] | W = 41** | r = −0.81 |
| Caregivers | RE vs EO | 11.05 (4.87) | 7.59 (3.19) | −3.46 [−4.67, −2.26] | W = 15** | r = −0.96 |

*Note:* **p < .001 & FDR q < .001; EO = eyes-open resting baseline; EC = eyes-closed resting; EX = storytelling; RE = listening. Values are relative alpha power averaged across the predefined 10-electrode sensor-level set: Fp1, Fp2, F3, F4, F8, T3, T4, Fz, Cz, and Pz. For normally distributed difference scores, paired-samples t-tests were used and effect sizes are reported as Cohen's dz. For non-normal difference scores, Wilcoxon signed-rank tests were used and effect sizes are reported as rank-based r. FDR q values were calculated using the Benjamini–Hochberg procedure.

**TABLE 3.** Associations between MoCA scores and alpha power at Cz.

| Condition | n | Spearman's ρ | 95% CI | p | FDR q |
|---|---|---|---|---|---|
| Eyes-closed resting | 64 | .13 | [−.12, .36] | .315 | .621 |
| Eyes-open resting | 64 | −.27 | [−.48, −.03] | .031 | .125 |
| Storytelling | 64 | −.08 | [−.32, .17] | .520 | .621 |
| Listening | 64 | −.06 | [−.30, .19] | .621 | .621 |

*Note:* MoCA = Montreal Cognitive Assessment; FDR = false discovery rate. Because MoCA scores were not normally distributed, Spearman's rank correlation coefficients were used. FDR-adjusted q values were calculated across the four condition-specific MoCA–Cz alpha correlations. No association survived FDR correction.

**TABLE 4.** Alpha-band inter-brain similarity and within-site surrogate analysis.

| Condition | Observed Fisher's z, M (SD) | Surrogate Fisher's z, M (SD) | 95% surrogate interval | Permutation p | FDR q |
|---|---|---|---|---|---|
| Eyes-closed resting | 0.62 (0.49) | 0.55 (0.06) | 0.44, 0.67 | .112 | .112 |
| Eyes-open resting | 0.80 (0.46) | 0.72 (0.05) | 0.63, 0.81 | .049 | .092 |
| Storytelling | 1.00 (0.48) | 0.93 (0.04) | 0.85, 1.01 | .041 | .092 |
| Listening | 0.94 (0.66) | 0.86 (0.05) | 0.77, 0.96 | .069 | .092 |

*Note:* Inter-brain similarity was calculated using Pearson spatial correlations between older adults' and caregivers' alpha-power patterns across the predefined 10-electrode set and transformed using Fisher's z. Surrogate distributions were generated by randomly re-pairing older adults and caregivers within the same museum site across 10,000 permutations. Permutation p values are one-tailed tests of whether observed dyad-level similarity exceeded the surrogate distribution. FDR q values were calculated across the four condition-specific surrogate comparisons.
